# Genetics of cardiometabolic disease progression

**DOI:** 10.1101/2025.02.01.25321518

**Authors:** Johanne M. Justesen, Guhan Venkataraman, Yosuke Tanigawa, Ruilin Li, Trevor Hastie, Robert Tibshirani, Joshua W. Knowles, Manuel A. Rivas

**Affiliations:** Department of Biomedical Data Science, Stanford University, CA, United States; Department of Medicine, Cardiovascular Institute, Diabetes Research Center, Prevention Research Center, Stanford University, CA, United States; Institute for Computational and Mathematical Engineering, Stanford University, Stanford, 94305, United States; Department of Statistics, Stanford University, CA, United States

**Keywords:** Genetics, cardiometabolic diseases, polygenic hazards score, Lasso, prediction, hyperlipidemia, operation, disease progression, lipids, lasso, Cox model

## Abstract

**Background:** Genome-wide association studies have been crucial in gaining insights into the genetics of cardiometabolic diseases. However, little is known about the genetics of cardiometabolic disease progression which may have both a different genetic architecture and significant implications for treatment decisions. Disease progression can be ascertained by the time from the first disease diagnosis to a second qualifying event (e.g. diagnostic lab, code or procedure). While data of this nature have been available in large repositories such as the UK Biobank, large-scale genome-wide screens in a time-to-event setting have been extremely challenging due to various computational and statistical challenges.

**Methods and Results:** We applied our method, snpnet-Cox, that has proven to be an effective method for simultaneous variable selection and estimation in high-dimensional settings, to examine the genetic contributions to cardiometabolic disease progression, measured by time from disease diagnosis to time of complication/comorbidity diagnosed or procedure in the UK Biobank. We apply a Cox regression model in a time-to-event setting to compute polygenic hazard scores (PHS). We identified ten new PHS that significantly predict disease progression. One example is the PHS that significantly predicts the time from hyperlipidemia diagnosis to having coronary artery bypass graft (CABG) surgery performed (Hazards Ratio 1.3 per PHS standard deviation: *p*=4.5×10^-9^). In this PHS, we identified a common variant, rs11041816 (downstream of *LMO1*), which protects against this disease progression (beta = −0.05).

**Conclusion:** snpnet-Cox is a fast and reliable tool to compute PHS capturing genetics in the time-to-event setting. The computed PHS can be used to stratify individuals with an underlying diagnosis (e.g. hyperlipidemia) into different trajectories disease progression (e.g CABG) thereby identifying potential points of intervention. With more time-to-event data to be released, this approach can provide great insight into disease progression at the fraction of computational cost necessary. We make available ten polygenic hazard scores that we find to be significant predictors of cardiometabolic disease progression.

## Introduction

Cardiometabolic diseases are an increasing global health burden affecting about a third of the world population ^1–2^. These common, chronic metabolic conditions such as cardiovascular disease (including hypertension, dyslipidemia, atherosclerosis and heart failure) and type 2 diabetes are a major source of morbidity and mortality but are highly heterogeneous in disease progression. There are many well-known clinical modifiers of disease progression (e.g smoking for atherosclerosis and obesity for type 2 diabetes)^2^. Similarly, genome-wide association studies (GWAS) have identified thousands of SNPs that are associated with incident and prevalent cardiometabolic disease status^3^. However, the genetic factors that alter the course of disease progression are not well understood. Furthermore, it is unclear whether the genetic risk factors that alter initial disease presentation are the same that alter disease progression. From a clinical perspective, it would be highly relevant to identify individuals who are genetically predisposed to a specific prognosis and therefore could particularly benefit from targeted interventions.

Indeed, a key prerequisite for precision health and medicine is a deeper understanding of disease state progression. Populations in biobanks with longitudinal medical record data are useful in this regard. For instance, one measure of disease progression is time from initial disease diagnosis to a comorbidity diagnosis (i.e., time from first disease diagnosis of type 2 diabetes to the diagnosis of a relevant comorbidity like diabetic neuropathy). Another measure of progression reflecting disease severity is captured by the disease-relevant procedures performed after the initial diagnosis (i.e. time from initial diagnosis to subsequent surgery, as recorded by Office of Population Censuses and Surveys (OPCS) Classification of Interventions and Procedures version 4 (OPCS-4).

The UK Biobank provides an opportunity to study genetics in longitudinal settings, providing times of disease diagnoses as well as dates of surgical procedures. This cohort has genotypic data on about 500,000 individuals coupled with unique phenotype information from electronic health records (EHRs) comprising primary care, hospital inpatient, cancer registry and death records ^4^.

Statistical models for disease progression have been challenging to develop. The Cox proportional hazards model provides a flexible mathematical framework to describe the relationship between the time to an event and various independent variable features, allowing for the calculation of a time-dependent baseline hazard useful in modeling disease progression. This model is regularly used to analyze time-to-event data in prospective epidemiological settings. However, these survival models face computational and statistical challenges when the predictors are ultra-high dimensional (i.e. feature dimension is far greater than the number of observations) and in large-scale settings where the data exceeds memory limits. To overcome this issue, we have previously developed a batch-screening iterative lasso (BASIL) algorithm to fit a Cox proportional hazard model by maximizing the Lasso partial likelihood function (snpnet-Cox)^5^. The Lasso is an effective tool for high-dimensional variable selection and prediction ^6^. We have previously applied snpnet-Cox to examine genetics of common complex diseases in a time-to-event setting ^7^.

Here, we apply snpnet-Cox to compute polygenic hazard scores (PHS) that examine the genetic variation of cardiometabolic disease progression in white British individuals from the UK Biobank (n = 337,129). Disease progression is defined as time from disease diagnosis to either next recorded disease or undergoing operational procedure. We replicate our findings using these PHS in a separate, non-British white population in UK Biobank (n = 28,134). The output of our algorithm is the full Lasso path, the parameter estimates at all predefined regularization parameters, as well as their validation accuracy measured using the concordance index (C-index) (Figure 1). We show that ten PHS significantly predict disease progression. Several of the PHS variants are known from GWAS to be associated with cardiometabolic disease status. However, we identify additional variants such as a common variant downstream of *LMO1* (rs11041816) that protects against progression from a hyperlipidemia diagnosis to undergoing coronary artery bypass graft (CABG).

**Figure 1:**
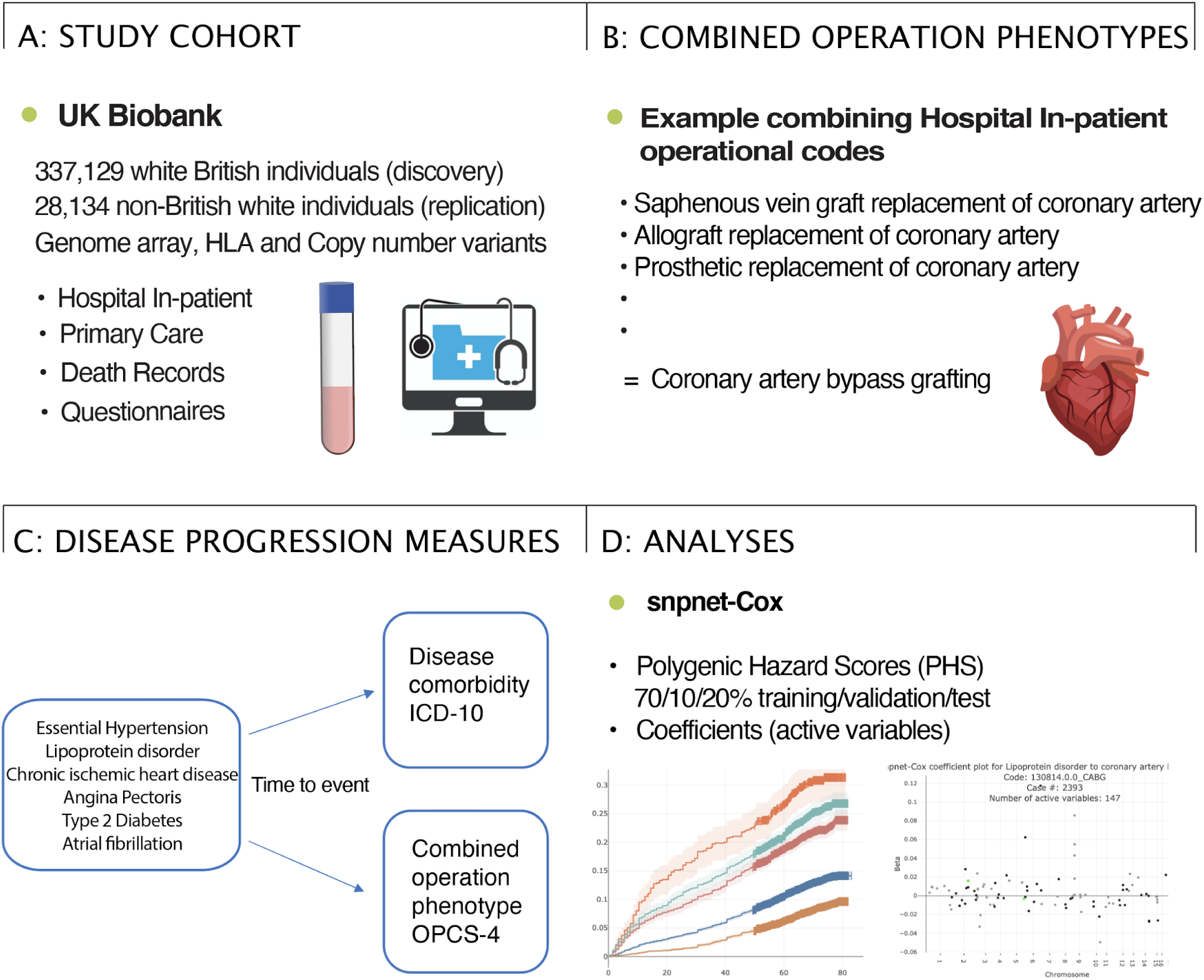
Study overview. A: Summary of the UK Biobank genotype and phenotype data used in the study. We used the white British subset of the UK Biobank for discovery and the non-British white subset for replication, analyzing LD-pruned and quality-controlled variants in relation to cardiometabolic disease progression measures based on electronic health records (hospital in-patient, primary care, and death records) and questionnaires with date of event. B: We created cardiometabolic surgical phenotypes by combining in-hospital operational codes from the OPCS-4 (Supplementary Table 1). C: We study the genetics of disease progression in a time-to-event setting in which disease progression is measured by time from disease diagnosis (International Classification of Diseases (ICD)-10 code) to disease comorbidity (next ICD-10 code) or having operation performed (next OPCS-4 code). D: We applied our R package, snpnet-Cox, to fit the Cox proportional Hazard model on the large genotype-disease progression dataset and compute polygenic hazard scores (PHS). The output of our algorithm is the full Lasso path.

## Methods

### Study population

The UK Biobank is a large, prospective population-based cohort study including individuals collected from multiple sites across the United Kingdom ^4,8^. It contains extensive genotypic and phenotypic information for 500,000 individuals aged 40-69 years when recruited in 2006-2010, including genome-wide genotyping, questionnaires, and physical measures for a wide range of health-related outcomes. In addition, information has been linked to registries from primary care, in-hospital records, death records, and the cancer registry. The discovery analyses were performed in white British individuals (n=337,129), and the replication in non-British white individuals (n=28,134).

### Genetic data preparation

We used genotype data from the UK Biobank dataset release version 2 and the hg19 human genome reference for all analyses in the study. To minimize the variabilities due to population structure in our dataset, we restricted our discovery analyses to include 337,129 unrelated white British individuals. In replication analyses, we included 28,134 non-British white individuals. Focusing on the entire study population of 337,129 individuals and 147,604 variants that are marked as “in_PCA" in the variant QC file (ukb_snp_qc.txt) from UK Biobank Sudlow et al. (2015)^4^, we compute the principal components of genetic variants using “–pcaapprox" sub-command in PLINK2 ^9^. We use the top 10 principal components, sex genotyping array and age at initial disease diagnosis as covariates.

The genetic variants used in this study were a combination of the directly genotyped variants from the UK Biobank (release version 2)^4^, the imputed allelotypes in human leukocyte antigen (HLA) allelotypes ^10^, and copy number variations (CNVs)^11^, resulting in a genotype matrix of 1,080,968 variants, as described in Sinnot-Armstrong et al 2021^12^. As a preprocessing step, we excluded variants with a missing rate greater than 10% and variants whose minor allele frequency is less than 0.001, which left approximately 700,000 variants remaining as features for our progression analyses.

### Disease phenotypes ICD-10

Time-to-event phenotypes were derived from First Occurrence of Health Outcomes data as defined by 3-character ICD-10 codes in UK Biobank’s Category 1712. The First Occurrence data-fields were generated by combining: read code information in the primary care data (Category 3000); ICD-9 and ICD-10 codes in hospital inpatient data (Category 2000); ICD-10 codes in death registry records (Field 40001, Field 40002); and self-reported medical condition codes (Field 20002), reported at baseline or subsequent UK Biobank assessment center visits as 3-character ICD-10 codes (censoring date March 1st 2020). A group of ʻSpecial event dates’ from primary care data were changed similarly to UK Biobank self-reported events.

- 1902-02-02 was changed to date of birth (total of 831 events)
- 1903-03-03 was changed to date of birth + 6 months (total of 651 events)
- 2037-07-07 is a date in the future and was removed from analysis (total 6 events)

Algorithmically-defined outcomes (based on data from Category 42) include phenotypes of select health-related events obtained through algorithmic combinations of coded information from the UK Biobank’s baseline assessment data collection. The data were derived from self-reported medical conditions, operations and medications together with linked data from hospital admissions and death registries (censoring date March 1st 2019).

- 334 items have date of unknown value, 1900-01-01, and were removed from analyses

To calculate age at disease diagnosis, death, and censoring, we computed the dates of birth (DOB) using the Month of Birth Data Field (Data-Field 52) and Year of Birth (Data-Field 34). All DOB were set to the first day of their birth month to avoid negative age of disease values. All ages at events = 0 were changed to the age event of 1 month, since snpnet-Cox uses values greater than 0. There were 40 dates of events that were one month before birth that were changed to DOB.

Data was structured by an *n* by *p* matrix of covariate values, where each row corresponded to an individual from the UK Biobank and each column a covariate. *y* was an *n-*length vector of event/death/censoring times, and status was an *n-*length vector where 0 was assigned if the entry in *y* was indicative of right censoring (i.e. the event had not yet happened at the time the data was collected) or death, and 1 was assigned if the event occurred.

### Phenotypes generated from surgical procedures OPCS-4

In the UK, OPCS Classification of Interventions and Procedures version 4.7 is the system used to code interventions, and ICD-10 the system for diagnoses. Each admission may contain several episodes, each corresponding to the care provided during a hospitalization. Using operational procedure codes (OPCS-4), we constructed phenotypes for common medical procedures for cardiometabolic conditions and complications. Most codes relevant to cardiac surgery belong to OPCS chapters K (Heart) and L (Arteries and veins). Multiple OPCS-4 codes were combined to create the phenotypes created, which include those described in Supplementary table 1. The two surgical phenotypes with the highest number of cases were percutaneous coronary intervention (angioplasty) and coronary artery bypass grafting (CABG).

Percutaneous coronary intervention (PCI, formerly known as coronary angioplasty and stent implantation) is a procedure that improves blood flow to the heart and thereby decreases heart-related chest pain (angina). By using a catheter (thin flexible tube) to place a small structure called a stent, it opens up blood vessels in the heart that have been narrowed by plaque buildup (a condition known as atherosclerosis).

Coronary artery bypass grafting (CABG) is an open chest procedure to perform direct revascularization of the heart by using a suitable vein from the chest, arm or leg for grafting to the coronary artery, thereby allowing the blood to bypass narrowings or blockages in the artery and reducing angina.

### Cardiometabolic disease progression phenotypes

We selected seven common cardiometabolic disease phenotypes with more than 15,000 cases in the UK Biobank to use as baseline diseases - Essential hypertension (ICD10: I10), lipoprotein disorder (E78 - hereafter we refer to lipoprotein disorder as hyperlipidemia given common clinical usage), chronic ischemic heart disease (I25), angina pectoris (I20), obesity (), non-insulin-dependent diabetes (type 2 diabetes) (E11) and atrial fibrillation (I48) (Supplementary Table 2). For disease progression outcomes, we included 21 disease and 12 operation phenotype outcomes which had more than 400 cases (Supplementary Table 3 and 4).

### BASIL algorithm to fit a Cox Proportional Hazard model on genotype-disease progression time dataset

To compute PHS, we use an R package, snpnet, which is based on a batch-screening iterative lasso (BASIL) algorithm that fits the full lasso solution path for very large and high-dimensional datasets, method previously described in Qian et al 2020^5^ and a Cox model application described in Li et al 2021^7^.

Briefly, this method is particularly suitable for large-scale and high-dimensional data that does not fit entirely in memory. Loading our UK Biobank data matrix with 1.08 million variants into R takes around 2.4 Terabytes of memory, which exceeds the size of most typical machines’ RAM. The Lasso is an effective tool for high-dimensional variable selection and prediction ^6^. In each iteration, we are able to effectively prune out variables (genetic variants) that are not relevant to disease progression, thereby eventually determining a “path” (of selected variants) relevant to disease.

We apply snpnet-Cox to the time-to-event setting where we analyze time from disease diagnosis to comorbidity event. Individuals were only included in the analysis if the age at disease diagnosis was prior to age of comorbidity diagnosis / surgical procedure. We split the dataset into a 70% training, 10% validation and 20% held out test set and apply snpnet-Cox with 50 iterations.

First, we assessed the predictive power of the PHS on time-to-event in the individuals in the held-out test set, thereby obtaining a *p*-value for each disease progression analysis. Second, we computed the hazard ratio (HR) for the scale (standard deviation (SD) unit) within different threshold percentiles (top 1%, 5%, 10% and bottom 10% compared to the 40-60%). Third, we computed the concordance index (C-index), which is a validation accuracy measure ^13^.

## Results

The ten cardiometabolic disease progression analyses with PHS *p*-values < 0.01 are listed in Table 2. Among all disease progression analyses, we found that significantly predictive PHS included a range of active variables from 2 to up to 188, highlighting the sparse property of Lasso in the Cox model (Table 1). The genetic predictive accuracy, C-index, ranged from 0.53 to 0.58, and HR per SD of PHS, from 1.13 to 1.31 (Table 1).

**Table 1:** Significant polygenic hazards scores (*p* < 0.01) computed using snpnet-Cox for cardiometabolic disease progression in white British individuals from UK Biobank.

| Phenotypes | N cases | Active variables | Scale HR | Scale $p$-value | Scale C | Top 1 % HR ($p$-value) | Top 5% HR ($p$-value) | Top 10% HR ($p$-value) | Bottom 10% HR ($p$-value) |
| --- | --- | --- | --- | --- | --- | --- | --- | --- | --- |
| Hyperlipidemia to CABG | 2393 | 147 | 1.31 | $4.49 \times 10^{-9}$ | 0.58 | 2.60 (0.0019) | 1.61 (0.023) | 1.74 (0.0040) | 0.57 (0.01) |
| Hyperlipidemia to chronic ischaemic heart disease | 9197 | 27 | 1.13 | $3.02 \times 10^{-7}$ | 0.53 | 1.25 (0.31) | 1.25 (0.055) | 1.12 (0.32) | 0.79 (0.014) |
| Hypertension to angioplasty | 3075 | 126 | 1.22 | $2.03 \times 10^{-6}$ | 0.56 | 1.97 (0.025) | 1.08 (0.72) | 1.71 (0.0016) | 0.81 (0.21) |
| Hypertension to T2D | 5747 | 3 | 1.15 | $2.90 \times 10^{-6}$ | 0.54 | 1.13 (0.68) | 1.18 (0.27) | 1.31 (0.11) | 0.96 (0.76) |
| Hyperlipidemia to myocardial infarction | 4201 | 188 | 1.16 | $2.34 \times 10^{-5}$ | 0.54 | 1.78 (0.045) | 1.14 (0.045) | 1.33 (0.077) | 0.90 (0.47) |
| Hyperlipidemia to angina | 5728 | 2 | 1.13 | $3.92 \times 10^{-5}$ | 0.53 | | 1.51 (0.0006) | 0.92 (0.60) | 0.90 (0.26) |
| Hyperlipidemia to angioplasty | 3636 | 10 | 1.13 | $7.53 \times 10^{-4}$ | 0.54 | 1.30 (0.42) | 1.04 (0.86) | 1.35 (0.072) | 0.80 (0.15) |
| Chronic ischaemic heart disease to CABG | 3548 | 5 | 1.13 | 0.0019 | 0.53 |  | 1.12 (0.54) | 1.34 (0.085) | 0.85 (0.27) |
| Hypertension to CABG | 1978 | 13 | 1.16 | 0.0040 | 0.55 | 1.84 (0.12) | 1.05 (0.85) | 1.24 (0.36) | 0.67 (0.078) |
| Angina to CABG | 2648 | 6 | 1.13 | 0.0078 | 0.53 | 1.24 (0.61) | 1.04 (0.89) | 1.75 (0.0023) | 0.93 (0.70) |

Scaled values (HR, p-value and C-index) are per standard deviation unit. N cases is the outcome (disease or surgery) number of cases. HR: Hazards ratio, CABG: Coronary Artery bypass Graft. T2D: Type 2 diabetes

All results are provided on the Global Biobank Engine (GBE)’s snpnet-Cox Disease Progression application (https://biobankengine.shinyapps.io/disease_progression/).

### Polygenic hazard score predicts progression from hyperlipidemia to coronary artery bypass surgery

Hyperlipidemia (“Disorders of lipoprotein metabolism and other lipidemias” - ICD-10 code E78) is general code applied to disorders with a spectrum of abnormalities in the levels of blood lipids (primarily cholesterol and triglycerides) carried by lipoproteins (e.g. Low density lipoprotein cholesterol (LDL)-C, high density lipoprotein cholesterol HDL-C) in the blood. Over time, increased levels of lipids in the blood, particularly LDL-C, can result in buildup of plaque in, narrowing of, or in the worst cases, blockage of arterial blood vessels especially in the heart (coronary artery disease). While mild coronary artery disease can be effectively managed with medications alone, advanced coronary artery disease may require percutaneous coronary interventions (including angioplasty and stent placement) and, when especially severe, coronary artery bypass graft (CABG) surgery to restore blood flow to the heart^14^. Susceptibility to and progression of coronary artery disease in the setting of hyperlipidemia is highly heterogeneous and the heterogeneity is only partly explainable by standard clinical characteristics (e.g. age, absolute lipid levels, use of medications to lower lipid levels).

In the UK Biobank, we estimated a HR of 1.31 per SD of PHS (*p* = 4.49×10^-9^) for the disease progression from hyperlipidemia to CABG (Table 2). This PHS was composed of 147 active variables and had a C-index of 0.58 (Table 1).

For individuals in the top 1%, 5% and 10% of the PHS distribution compared to the 40-60%, we estimated a HR of 2.6, 1.6 and 1.7 respectively, whereas individuals in the bottom 10% of the PHS distribution had significantly lower risk of having CABG surgery, with a HR of 0.57 (*p* = 0.01) (Figure 2A and Supplementary table 5).

**Figure 2:**
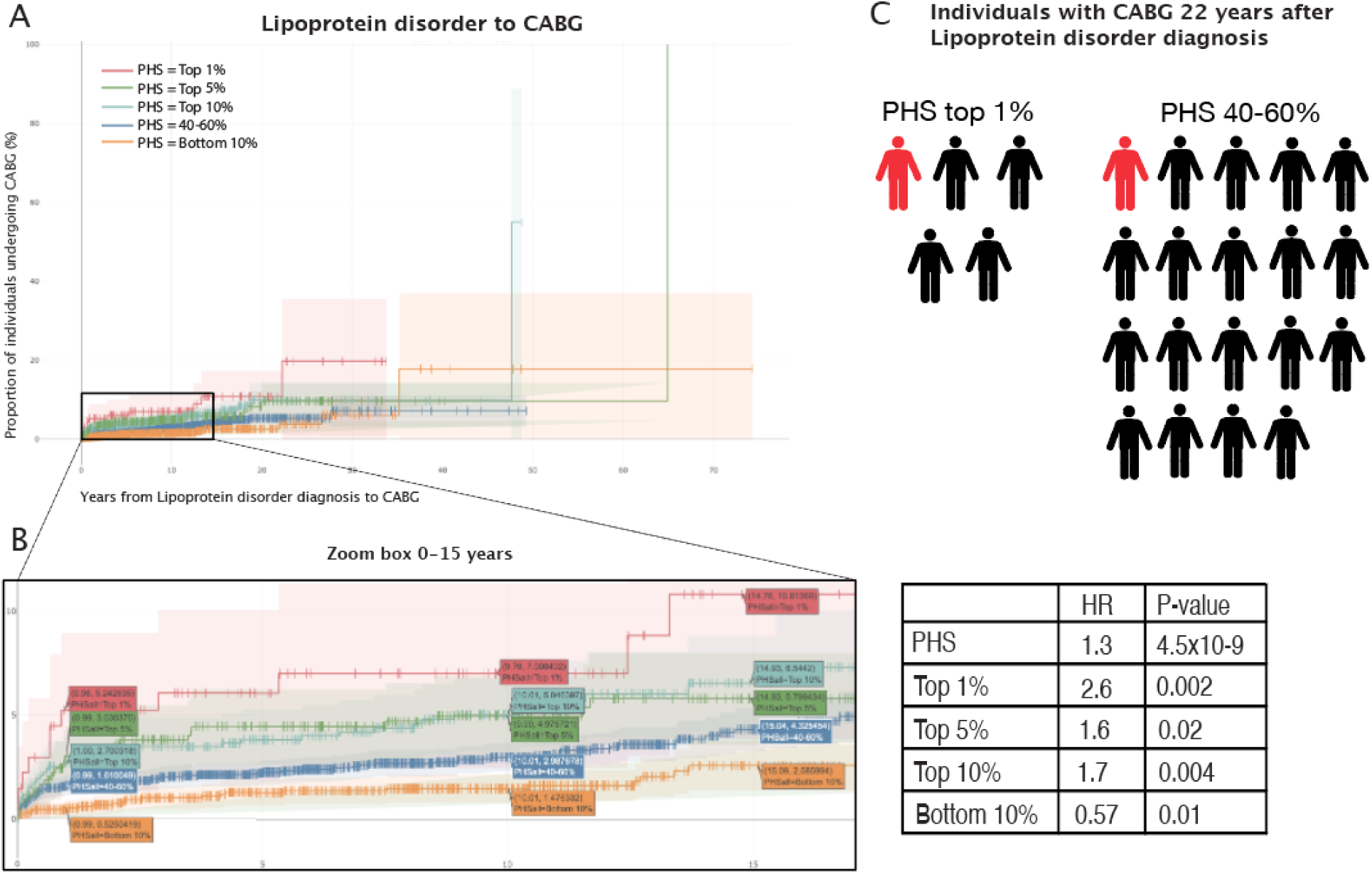
Kaplan-Meier curves and PHS extremes. A: Kaplain-Meier curves for percentiles of polygenic hazard scores (PHS) for variants selected by snpnet-Cox, in the held out test set (red - top 1%, green - top 5%, light blue - top 10%, blue - 40-60%, and orange bottom 10%: ticks represent censored observations). B: Zoom box highlighted are the proportion of coronary artery bypass graft (CABG) surgeries, 1, 10 and 15 years after hyperlipidemia diagnosis across the percentile scores. CABG case count = 2393. C: For the top 1% PHS the Hazards ratio of CABG.

Just one year after a hyperlipidemia diagnosis, we found that 5.24% of individuals in the top 1% of the PHS distribution had CABG performed, whereas only 0.53% of the bottom 10% and 1.61% of the 40-60 percentile of the PHS underwent CABG surgery (Figure 2B).

Further, we found the risk of having CABG fifteen years after hyperlipidemia diagnosis was more than doubled for individuals in the top 10% of the PHS distribution (6.54%) compared with the bottom 10% (2.59%) (Figure 2B).

In 2008, the global prevalence of increased plasma cholesterol levels was estimated to be ∼39% among individuals 25 years and older^15^ and more than one-third caused by CAD and ischemic stroke were attributable to increased plasma LDL-cholesterol levels ^16^. Compared with the general population, individuals in the UK Biobank are generally healthier due to the “healthy volunteer” selection bias ^17^; we find that 21% of all UK Biobank individuals have hyperlipidemia, with a mean age of 58 years at diagnosis. However, this condition is also becoming diagnosed more frequently earlier in life, and in the US, 20% of children (age 6-19 years) have adverse lipid levels^18^. In the UK Biobank, we find that 22 years after hyperlipidemia diagnosis, among the PHS distribution’s top 1%, around 1 in 5 (19.7 %) will have had CABG surgery performed. In contrast, for the bottom 10% and 40-60% of the PHS distribution, 1 in 26 (3.79%) and 1 in 19 (5.29%) will have had the CABG surgery, respectively (Figure 2A and 2C). This highlights the potential relevance of applying PHS in the context of screening patients for further followup examinations.

### Active variables of the polygenic hazards score for hyperlipidemia to coronary artery bypass surgery

We applied the BASIL algorithm to generate predictive models with sparse solutions using the genotype and phenotype data, thereby identifying the features (genetic variants) that are most relevant for disease progression in the time-to-event setting.

In the analysis of the progression progressing from hyperlipidemia to CABG, snpnet-Cox identified 147 genetic variants (active variables), of which several have been previously identified as risk loci from GWAS of Coronary artery disease, hyperlipidemia and related traits. We identified rs10455872 (MAF = 0.08), an intron variant in the *LPA*, to associate with an effect size (beta) of 0.12, i.e. 12% increased hazards of disease progression (Figure 3). This gene encodes lipoprotein(a), a large lipoprotein made by the liver, and is known to be an independent risk factor for cardiovascular diseases and causal of atherosclerosis, heart attacks, strokes, and heart failure ^19^. Among other known loci are *PHACTR1* (beta = 0.06 ^20^, *CDKN2B-AS1* (beta = 0.09) ^21–23^ and *ATP2B1* (beta = 0.02) ^24,25^. Additionally, we identified a missense variant, rs1990760, in *IFIH1*, to increase the risk of progressing to CABG with beta = 0.02 (Figure 3). This variant has previously been associated with reduced risk of hypothyroidism (OR 0.92 CI 0.90, 0.94; *p* = 9.3 × 10^−17^) and was suggested to influence coronary artery disease risk (OR 0.97 CI 0.96, 0.99; *p* = 2.5 × 10^−5^) ^26^. Furthermore, we found a common variant (MAF = 0.46) downstream of *LMO1*, rs11041816, which showed a protective effect of progression to having CABG performed, beta = −0.05. This gene encodes a transcriptional regulator and it has been found to regulate transcription by competitively binding to specific DNA-binding transcription factors ^27^. *LMO1* has previously been associated with other metabolic traits such as systolic and diastolic blood pressure ^28^, fasting blood glucose ^29^, body mass index ^30^ and birth weight ^31^ but not coronary artery disease.

**Figure 3.**
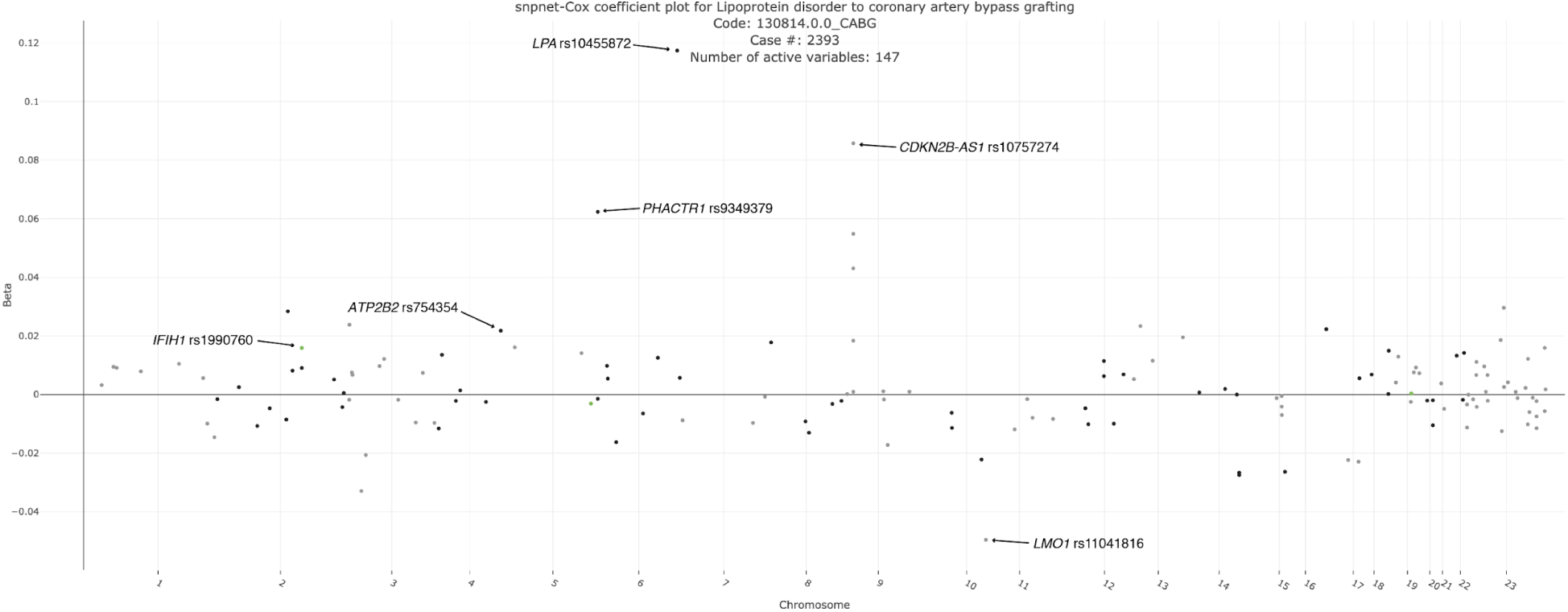
snpnet-Cox selected variables. Plot of snpnet-Cox coefficients in analysis of progression from hyperlipidemia to coronary artery bypass graft (CABG) surgery, with 147 active variables. Green dots represent protein-altering variants. https://biobankengine.shinyapps.io/disease_progression/

Phenome wide association study (PheWAS) was performed for rs11041816 using the Global Biobank Engine, we found association with vascular heart problems diagnosed by doctor - high blood pressure (https://biobankengine.stanford.edu/RIVAS_HG19/variant/11-8243798-A-G and Supplementary Table 6). Additionally, we found an independent variant rs4480535 upstream of *LMO1* protected from CABG in the Finngen study beta = −0.12 and *p* = 1.2×10^-6^ (http://r4.finngen.fi/region/I9_CABG/gene/LMO1).

#### Replication of significant PHS in non British white individuals of UK Biobank

Next, we examined the significant PHS in the non British white individuals from UK Biobank (n=28,134) where four of the ten PHS significantly replicate (*p* < 0.05, Supplementary Table 3). Due to the smaller size of this cohort, we had very few cases for some traits, affecting the significance of the other six PHS. Yet, for the six PHS that did not significantly replicate, we found the PHS had the same direction of effect as observed in the discovery analyses.

#### *TCF7L2* is the main driver of progression from hypertension to type 2 diabetes

In the UK Biobank, we estimated a HR of 1.15 per SD of PHS (*p* = 2.90×10^-6^) for the disease progression from hypertension to T2D. This PHS was composed of only 3 active variables and had a C-index of 0.54 (Table 1). *TCF7L2* rs7903146 has a beta of 0.136. It is known to influence insulin secretion and glucose production and is the main loci known in relation to T2D^32^.

#### Applicability snpnet-cox

When biobanks have longer follow up time and include more individuals these type of analyses will have more power to identify loci. Even though the PHS computed for hyperlipidemia to embolectomy was not significant, this PHS included interesting SNPs such as the GPR107 (3 prime UTR variant rs1306, beta −0.058) that had a protective effect for the disease progression. This has previously been … Additionally, an intron variant of TBC1D4 (rs517130, beta −0.028) did also protect from progressing from hyperlipidemia to embolectomy. TBC1D4 confers muscle insulin resistance and type 2 diabetes^33^.

Furthermore, the method could also be applied to study other disease progression patterns like those observed in cancer.

## Discussion

In this study, we applied the batch-screening iterative Lasso (BASIL) algorithm implemented in the R snpnet package to study the genetic architecture of disease progression by computing PHS in a time-to-event setting within the UK Biobank. The BASIL algorithm to fit a Cox proportional hazard model on our large-scale and high dimensional dataset to generate predictive models with sparse solutions, which means that most variants have no effect. The output of this method is a list of the genetic variants that associate with the progression measure of interest. We generated a PHS using these genetic variants, identifying ten PHS that were significantly associated with disease progression in a white British cohort, of which four were replicated in a smaller, non-British white cohort. All results from snpnet-Cox have been made available in our Global Biobank Engine (https://biobankengine.shinyapps.io/disease_progression/) ^34^.

First, we assessed the predictive power of the PHS on time-to-event in the individuals in the held-out test set, thereby obtaining a *p*-value for each disease progression analysis. Second, we computed the hazard ratio (HR) for the scale (standard deviation (SD) unit) within different threshold percentiles (top 1%, 5%, 10% and bottom 10% compared to the 40-60%). Third, we computed the concordance index (C-index), which is a validation accuracy measure ^13^.

The best predictive PHS model (HR: 1.31, *p* = 4.49×10^-9^) was obtained for the severity progression from hyperlipidemia to CABG. In this process, hyperlipidemia causes coronary artery disease, which eventually progresses to a stage where surgery is needed to restore blood flow to the heart. Several of the variants are known from GWAS to associate with CAD. However, we did also identify a common variant downstream of *LMO1* which protects from this severity progression to CABG. This variant has not been described in relation to CAD from GWAS (NHGRI GWAS Catalog).

Applying BASIL algorithm we computed PHS for disease progression which importantly takes time to event into account. Prior studies have examined disease progression by recurrent events of MI and revascularization where a PRS based on risk loci from GWAS is associated with risk of subsequent events ^35–38^. However, variants that influence disease onset may not necessarily influence disease progression. By computing As we move towards whole genome sequencing, this algorithm is extremely fast and can handle the larger-than-memory datasets.

For cardiometabolic diseases the main determinant of patient well-being is not the diagnosis itself but instead the progression to a variety of complications. This varies substantially between patients. This is most likely due to a combination of lifestyle factors together with some unknown degree of genetic predisposition. However, in this setting, we cannot take either medication, physical activity or diet into account. We applied phenotypes which mainly are derived from linking to electronic health records, and the UK Biobank lifestyle measures are mainly from enrollment in this study. Additionally, we had the ICD-10 in 3-character codes and not the more specific 4-character codes.

A substantial challenge in detecting loci that associate with disease progression is sufficient sample size. Overall, we observed that for the ten PHS which significantly predicted cardiometabolic disease progression in the test sets, eight out of the ten were for the two phenotypes having most cases at baseline - hypertension (n=83,727) and hyperlipidemia (n=68,477). These exploratory settings may be underpowered for more of the cardiometabolic disease progression analyses. However, with even more time-to-event data coming online as biobanks continue to gather follow-up data, as well as the growing number of biobanks becoming available, this method can prove useful, especially with its ability to handle out-of-memory datasets.

For some traits, there are too few cases. However, even with a small number of cases the snpnet-Cox is able to select variables that are associated with disease, albeit with less confidence (as evidenced by the replication cohort’s inability to confirm all the PHS selected in the discovery cohort).

Genetics could be a part of precision medicine adding to the traditional risk factors for disease. Potentially, using PHS for individuals who early in life have a cardiometabolic diagnosis, could help. Previously, other studies have computed polygenic scores from GWAS and tested in a time-to-event setting^39–40^, whereas here we take advantage of computing the PHS in the time-to-event setting.

In the usual GWAS setting, cases are defined as individuals who have the disease or trait of interest at any time point recorded. However, taking into account time-at-diagnosis, and, in this case, time for disease to progress, is relevant since this time length is highly variable between patients. With more biobanks having the opportunity to link data to electronic health records, it will become increasingly important to develop methods and statistical approaches that can take advantage of the longitudinal nature of these observations. This could potentially facilitate improved prediction, drug development and thereby better precision medicine^41^.

In conclusion, we applied a batch-screening iterative Lasso (BASIL) algorithm to find the lasso path of Cox proportional hazards models to study disease progression using genotype-phenotype dataset from UK Biobank. We selected 7 common complex cardiometabolic traits to identify genetic variants that are associated with time-to-event disease and surgical outcomes. We find 10 of these computed PHS that predict cardiometabolic disease progression. The genetics of disease status and disease progression overlap. However, we do identify putative genetic markers potentially important for disease progression such as the variant in *LMO1* which protects from progressing from a hyperlipidemia to CABG surgery. Future studies to illuminate the biological mechanism of this gene.

## Data Availability

All data is available from UK Biobank. The results are publicly available at https://biobankengine.shinyapps.io/disease_progression/

## Acknowledgements and funding sources

This research has been conducted using the UK Biobank Resource under Application Number 24983, “Generating effective therapeutic hypotheses from genomic and hospital linkage data” (http://www.ukbiobank.ac.uk/wp-content/uploads/2017/06/24983-Dr-Manuel-Rivas.pdf). We thank all of the participants in the UK Biobank study. This work was supported by the National Human Genome Research Institute (NHGRI) of the National Institutes of Health (NIH) under award R01HG010140. The content is solely the responsibility of the authors and does not necessarily represent the official views of the National Institutes of Health. Some of the computing for this project was performed on the Sherlock cluster. We would like to thank Stanford University and the Stanford Research Computing Center for providing computational resources and support that contributed to these research results.

J.M.J. is supported by grant NNF17OC0025806 from the Novo Nordisk Foundation and the Stanford Bio-X Program. J.W.K is supported by NIH grants: U41HG009649, R01 DK116750, R01 DK120565, P30DK116074. M.A.R. is supported by Stanford University and a National Institute of Health center for Multi- and Trans-ethnic Mapping of Mendelian and Complex Diseases grant (5U01 HG009080).

## Author Contributions

JMJ, GV, YT, RL, MA did quality control and analyses in UKB. Combining new phenotypes was performed by JMJ, JWK and MA. Statistical analyses and methods were evaluated by JMJ, GV, YT, RL, TH, RT and MA. The manuscript was written by J.M.J and M.A.R. and revised by all the co-authors. All co-authors have approved of the final version of the manuscript.

## Supplementary Materials

Phenotype definitions: https://docs.google.com/spreadsheets/d/1viEa4V5Se-czkUca_i_dsaH17vSMQkfJtTbnKl_xRIQ/edit#gid=0

## Supplementary tables

**Supplementary Table 1.** Overview of surgical phenotypes generated from multiple OPCS-4 codes from in-hospital records in UK Biobank.

| Name | Characteristics | OPCS-4 codes |
| --- | --- | --- |
| Percutaneous Coronary intervention (angioplasty) |  | K491 K492 K493 K494 K498 K499 K501<br>K502 K503 K504 K508 K509 K751 K752<br>K753 K754 K758 K759 |
| Coronary artery bypass grafting (CABG) | A procedure to improve poor blood flow to the heart. CABG uses blood vessels from another part of the body and connects them to blood vessels above and below the narrowed artery, bypassing the narrowed or blocked coronary arteries. | K401 K402 K403 K404 K408 K409 K411<br>K412 K413 K414 K418 K422 K424 K431<br>K433 K439 K441 K442 K448 K449 K451<br>K452 K453 K454 K455 K456 K458 K459<br>K461 K462 K463 K464 K469 |
| Carotid artery surgery |  | L291 L293 L294 L295 L297 L298 L299 L301<br>L302 L303 L304 L305 L308 L309 L311 L313<br>L314 L318 L319 O121 O122 O128 O129 |
| Abdominal aortic aneurysm |  | L181 L182 L183 L184 L185 L186 L188 L189<br>L191 L192 L193 L194 L195 L196 L198 L199<br>L271 L272 L273 L274 L275 L276 L278 L279<br>L281 L282 L283 L284 L285 L286 L288 L289 |
| Femoral/Iliac aneurysm |  | L491 L492 L493 L495 L496 L498 L499<br>L511 L512 L513 L514 L515 L516 L518<br>L519 L521 L522 L528 L531 L532 L533<br>L538 L539 L541 L542 L544 L548 L549<br>L571 L572 L573 L575 L591 L592 L593<br>L594 L595 L596 L597 L598 L599 L601<br>L602 L603 L604 L608 L609 L622 L623<br>L624 L628 L629 L631 L632 L635 L638<br>L639 |
| Embolectomy | Removal of blood clot | L124 L131 L253 L263 L303 L343 L383 L392<br>L421 L432 L461 L532 L542 L622 L632 L701<br>L712 |
| Peripheral artery interventions |  | L181 L182 L183 L184 L185 L186 L188 L189<br>L191 L192 L193 L194 L195 L196 L198 L199<br>L201 L204 L205 L206 L208 L211 L212 L213<br>L215 L216 L218 L231 L232 L233 L234 L235<br>L236 L238 L239 L251 L252 L253 L254 L255<br>L258 L259 L261 L262 L265 L266 L267 L268<br>L269 L271 L272 L273 L274 L275 L276 L278<br>L279 L281 L282 L283 L284 L285 L286 L288<br>L289 L291 L293 L294 L295 L297 L298 L299<br>L301 L302 L303 L304 L305 L308 L309 L311<br>L313 L314 L318 L319 L371 L373 L378 L381<br>L382 L383 L384 L388 L412 L413 L416 L418<br>L419 L423 L424 L428 L431 L432 L435 L438<br>L439 L451 L452 L463 L464 L468 L471 L474<br>L478 L491 L492 L493 L495 L496 L498 L499<br>L501 L503 L505 L506 L508 L509 L511 L512<br>L513 L514 L515 L516 L518 L519 L521 L522 |
|  |  | L528 L531 L532 L533 L538 L539 L541 L542<br>L544 L548 L549 L561 L562 L563 L571 L572<br>L573 L575 L578 L581 L582 L583 L584 L585<br>L587 L588 L589 L591 L592 L593 L594 L595<br>L596 L597 L598 L599 L601 L602 L603 L604<br>L608 L609 L622 L623 L624 L628 L629 L631<br>L632 L635 L638 L639 L661 L662 L663 L664<br>L665 L667 L701 L702 L704 L705 L708 L709<br>L711 L712 L714 L715 L716 L717 L718 |
| Pacemaker | A pacemaker (artificial pacemaker surgery) is a small device that is placed under the chest skin below the collarbone. One or two wires connect the pacemaker to the heart chambers. It generates a small electrical current that | K601 K605 K606 K607 K611 K615 K616<br>K617 K618 K619 |
|  | stimulates the heart muscle and ensures regular pumping activity. |  |
| Diabetes related eye surgery |  | C821 Y084 |
| Amputation | Amputation of leg, foot or toe | X091 X092 X093 X094 X095 X098 X101<br>X102 X103 X104 X108 X109 X111 X112<br>X118 X119 |
| End stage kidney disease surgery |  | X401 X402 X403 X404 X405 X406 X408<br>X409 M011 M012 M013 M014 M015 M018<br>M019 |
| Liver surgery |  | J011 J012 J013 J015 J018 J019 J021 J022<br>J023 J024 J025 J026 J027 J028 J029 J031<br>J032 J033 J034 J035 J038 J039 |
| Cardioversion | Restore abnormal heart rhythm<br>(arrhythmia) by sending electrical<br>signals to the heart through electrodes | X501 X502 |
|  | placed on the chest. |  |
| Percutaneous radiofrequency catheter ablation for atrial fibrillation |  | K574 K621 K622 K623 |

All OPCS-4 are listed on the UK Biobank website: https://biobank.ctsu.ox.ac.uk/crystal/field.cgi?id=41272
https://www.nice.org.uk/Media/Default/About/what-we-do/NICE-guidance/NICE-interventional-procedures/OPCS-reports-September-2012.pdf

**Supplementary Table 2.** Number of white British and non-British white individuals having the initial disease outcome in UK Biobank.

| Disease phenotype outcomes - ICD-10 | White British cases | Non-British white cases |
| --- | --- | --- |
| Essential (primary) hypertension - I10 | 83,727 |  |
| Disorders of lipoprotein metabolism and other lipidemias (hyperlipidemia)- E78 | 68,477 |  |
| Chronic ischaemic heart disease - I25 | 25,378 |  |
| Obesity | 22667 |  |
| Angina pectoris - I20 | 20,542 |  |
| Non-insulin-dependent diabetes mellitus (T2D) - E11 | 20,475 |  |
| Atrial fibrillation and flutter - I48 | 17,166 |  |

**Supplementary table 3.** Phenotypes included as disease outcomes (secondary diagnosis) for white British individuals in UK Biobank.

| Phenotype | ICD-10 | White British cases |
| --- | --- | --- |
| Disease phenotype outcomes - ICD-10 |  |  |
| Essential (primary) hypertension | I10 | 83727 |
| Disorders of lipoprotein metabolism and other lipidemias | E78 | 68477 |
| Chronic ischaemic heart disease | I25 | 25378 |
| Obesity | E66 | 22667 |
| Angina pectoris | I20 | 20542 |
| Non-insulin-dependent diabetes mellitus (T2D) | E11 | 20475 |
| Atrial fibrillation and flutter | I48 | 17166 |
| Acute myocardial infarction | I21 | 12514 |
| Chronic renal failure | N18 | 11928 |
| Peripheral artery disease | I73 | 7014 |
| Heart failure | I50 | 6812 |
| Other cardiac arrhythmias | I49 | 6339 |
| Acute renal failure | N17 | 6259 |
| Stroke, not specified as haemorrhage or infarction | I64 | 5918 |
| Pulmonary embolism | I26 | 5890 |
| Other diseases of liver | K76 | 5839 |
| Cerebral infarction | I63 | 3907 |
| Acute pancreatitis | K85 | 2440 |
| Unspecified renal failure | N19 | 2034 |
| Aortic aneurysm and dissection | I71 | 1983 |
| Other disorders of arteries and arterioles | I77 | 1631 |
| Cardiomyopathy | I42 | 1572 |
| Atherosclerosis | I70 | 1481 |
| Ulcer of lower limb | L97 | 1453 |
| Other inflammatory liver diseases | K75 | 1309 |
| Hypertensive renal disease | I12 | 1289 |
| Fibrosis and cirrhosis of liver | K74 | 1192 |
| Cardiac arrest | I46 | 1178 |
| Intracerebral hemorrhage | I61 | 942 |
| Hepatic failure, not elsewhere classified | K72 | 472 |
| Hypertensive heart disease | I11 | 408 |

**Supplementary table 4.**
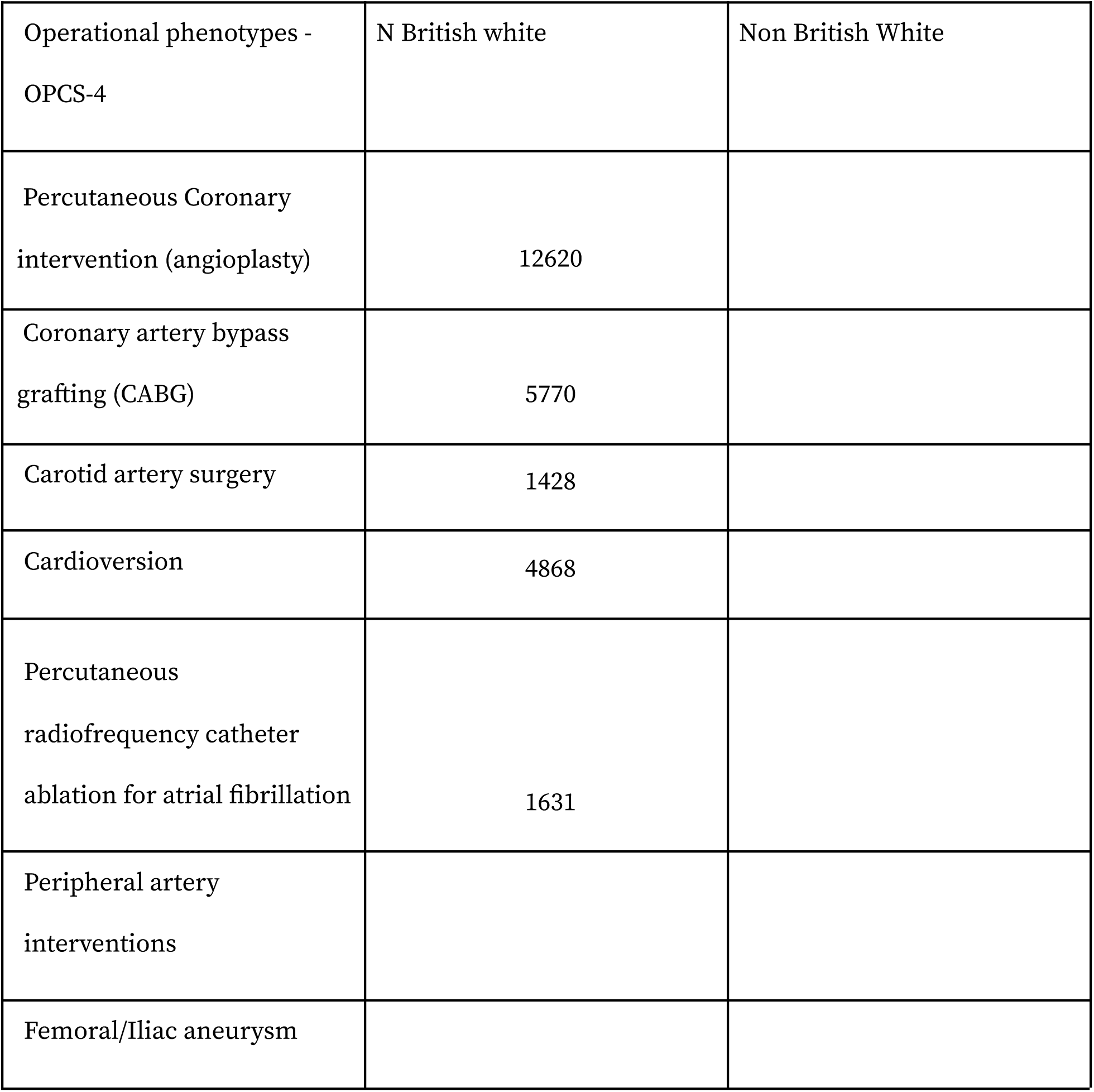
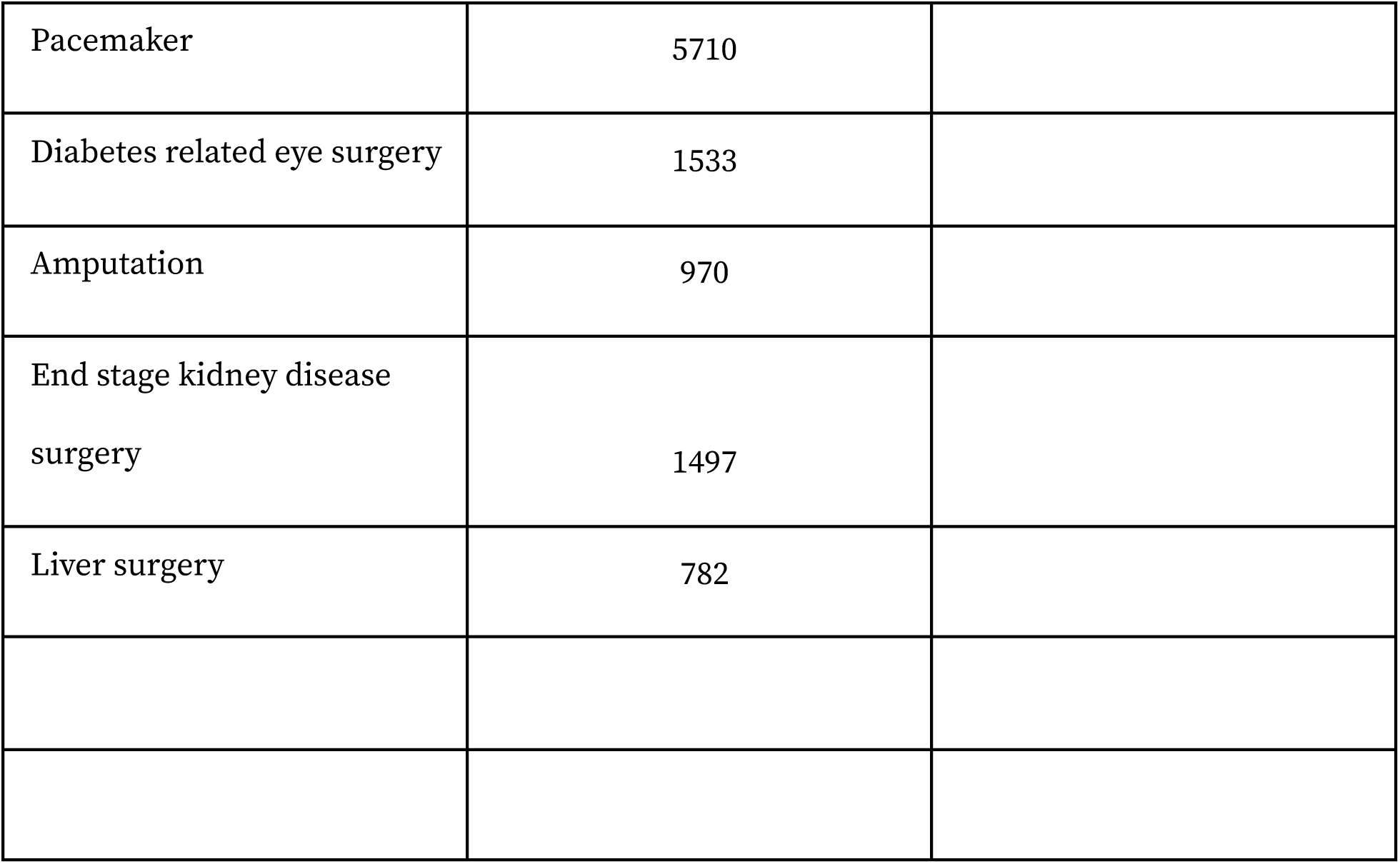
Phenotypes included as operation outcomes (surgeries performed after initial diagnosis) for white British individuals in UK Biobank.

**Supplementary Table 5.**
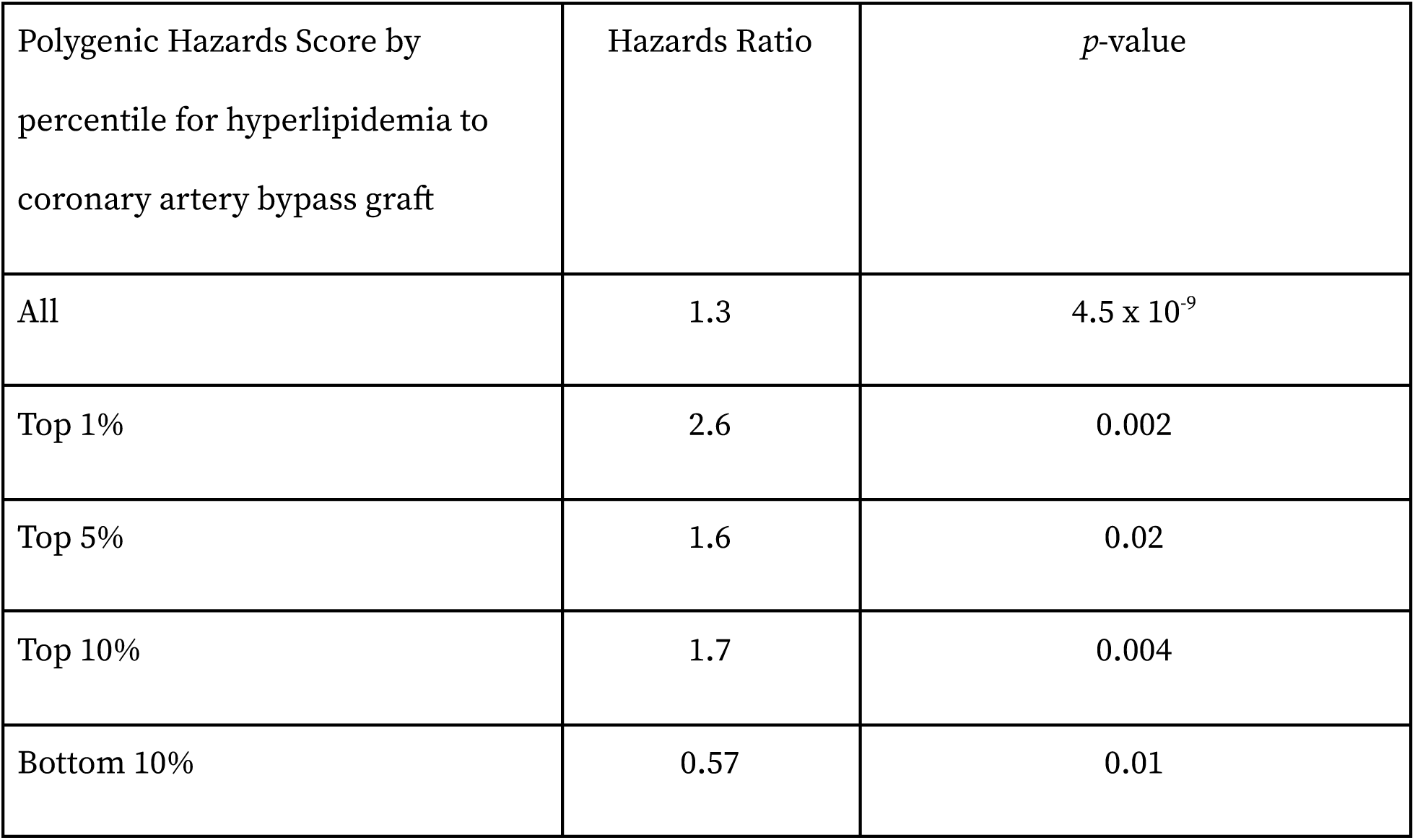
Time-to-event summary hazards ratio per standard deviation of the polygenic hazards score for all individuals and top 1, 5 and 10% and bottom 10% compared to 40-60%.

**Supplementary table 6.**
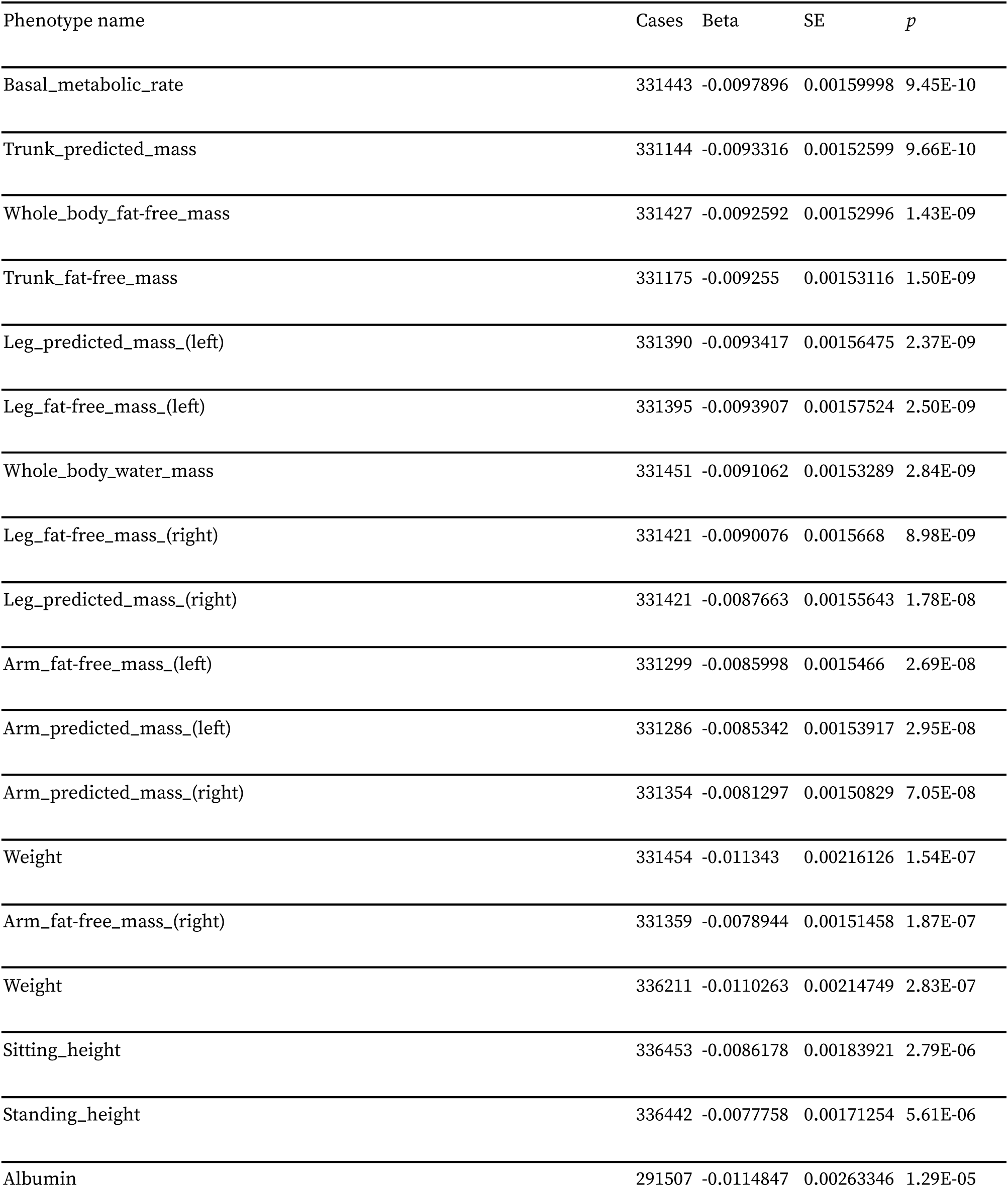
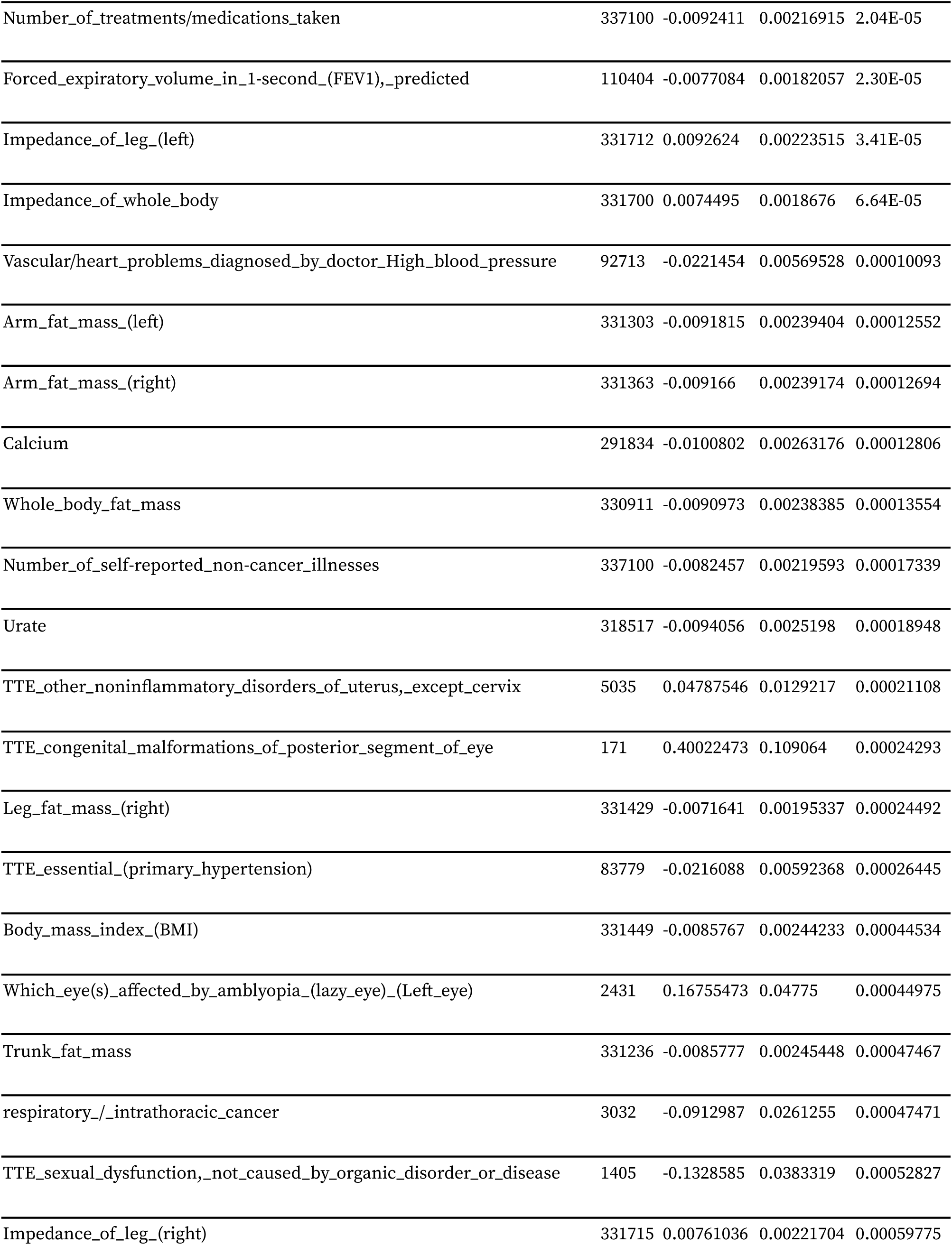
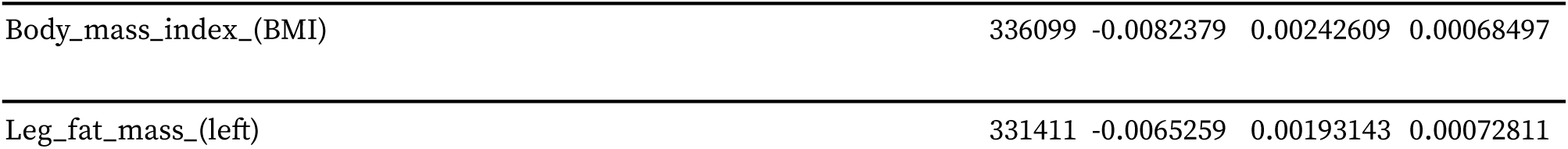
Phewas (*p* <= 1×10^-3^) for *LMO1* rs11041816 white British individuals from UK Biobank.

| Phenotype name | Cases | Beta | SE | *p* |
| --- | --- | --- | --- | --- |
| Basal_metabolic_rate | 331443 | -0.0097896 | 0.00159998 | 9.45E-10 |
| Trunk_predicted_mass | 331144 | -0.0093316 | 0.00152599 | 9.66E-10 |
| Whole_body_fat-free_mass | 331427 | -0.0092592 | 0.00152996 | 1.43E-09 |
| Trunk_fat-free_mass | 331175 | -0.009255 | 0.00153116 | 1.50E-09 |
| Leg_predicted_mass_(left) | 331390 | -0.0093417 | 0.00156475 | 2.37E-09 |
| Leg_fat-free_mass_(left) | 331395 | -0.0093907 | 0.00157524 | 2.50E-09 |
| Whole_body_water_mass | 331451 | -0.0091062 | 0.00153289 | 2.84E-09 |
| Leg_fat-free_mass_(right) | 331421 | -0.0090076 | 0.0015668 | 8.98E-09 |
| Leg_predicted_mass_(right) | 331421 | -0.0087663 | 0.00155643 | 1.78E-08 |
| Arm_fat-free_mass_(left) | 331299 | -0.0085998 | 0.0015466 | 2.69E-08 |
| Arm_predicted_mass_(left) | 331286 | -0.0085342 | 0.00153917 | 2.95E-08 |
| Arm_predicted_mass_(right) | 331354 | -0.0081297 | 0.00150829 | 7.05E-08 |
| Weight | 331454 | -0.011343 | 0.00216126 | 1.54E-07 |
| Arm_fat-free_mass_(right) | 331359 | -0.0078944 | 0.00151458 | 1.87E-07 |
| Weight | 336211 | -0.0110263 | 0.00214749 | 2.83E-07 |
| Sitting_height | 336453 | -0.0086178 | 0.00183921 | 2.79E-06 |
| Standing_height | 336442 | -0.0077758 | 0.00171254 | 5.61E-06 |
| Albumin | 291507 | -0.0114847 | 0.00263346 | 1.29E-05 |
| Number_of_treatments/medications_taken | 337100 | -0.0092411 | 0.00216915 | 2.04E-05 |
| Forced_expiratory_volume_in_1-second_(FEV1),_predicted | 110404 | -0.0077084 | 0.00182057 | 2.30E-05 |
| Impedance_of_leg_(left) | 331712 | 0.0092624 | 0.00223515 | 3.41E-05 |
| Impedance_of_whole_body | 331700 | 0.0074495 | 0.0018676 | 6.64E-05 |
| Vascular/heart_problems_diagnosed_by_doctor_High_blood_pressure | 92713 | -0.0221454 | 0.00569528 | 0.00010093 |
| Arm_fat_mass_(left) | 331303 | -0.0091815 | 0.00239404 | 0.00012552 |
| Arm_fat_mass_(right) | 331363 | -0.009166 | 0.00239174 | 0.00012694 |
| Calcium | 291834 | -0.0100802 | 0.00263176 | 0.00012806 |
| Whole_body_fat_mass | 330911 | -0.0090973 | 0.00238385 | 0.00013554 |
| Number_of_self-reported_non-cancer_illnesses | 337100 | -0.0082457 | 0.00219593 | 0.00017339 |
| Urate | 318517 | -0.0094056 | 0.0025198 | 0.00018948 |
| TTE_other_noninflammatory_disorders_of_uterus,_except_cervix | 5035 | 0.04787546 | 0.0129217 | 0.00021108 |
| TTE_congenital_malformations_of_posterior_segment_of_eye | 171 | 0.40022473 | 0.109064 | 0.00024293 |
| Leg_fat_mass_(right) | 331429 | -0.0071641 | 0.00195337 | 0.00024492 |
| TTE_essential_(primary_hypertension) | 83779 | -0.0216088 | 0.00592368 | 0.00026445 |
| Body_mass_index_(BMI) | 331449 | -0.0085767 | 0.00244233 | 0.00044534 |
| Which_eye(s)_affected_by_amblyopia_(lazy_eye)_(Left_eye) | 2431 | 0.16755473 | 0.04775 | 0.00044975 |
| Trunk_fat_mass | 331236 | -0.0085777 | 0.00245448 | 0.00047467 |
| respiratory/_intrathoracic_cancer | 3032 | -0.0912987 | 0.0261255 | 0.00047471 |
| TTE_sexual_dysfunction,_not_caused_by_organic_disorder_or_disease | 1405 | -0.1328585 | 0.0383319 | 0.00052827 |
| Impedance_of_leg_(right) | 331715 | 0.00761036 | 0.00221704 | 0.00059775 |
| Body_mass_index_(BMI) | 336099 | -0.0082379 | 0.00242609 | 0.00068497 |
| Leg_fat_mass_(left) | 331411 | -0.0065259 | 0.00193143 | 0.00072811 |

**Supplementary Table 7.** Replication of significant PHS associations in non-British White individuals from UK Biobank (n=28,134)

| Phenotypes | N cases | Scale HR | Scale P-value | Scale C |
| --- | --- | --- | --- | --- |
| Hyperlipidemia to CABG | 145 | 1.15 | 0.09 | 0.54 |
| Hyperlipidemia to chronic ischaemic heart disease | 595 | 1.16 | $2.8 \times 10^{-4}$ | 0.55 |
| Hypertension to angioplasty | 201 | 1.10 | 0.19 | 0.53 |
| Hypertension to T2D | 381 | 1.21 | $2.40 \times 10^{-4}$ | 0.55 |
| Hyperlipidemia to myocardial infarction | 276 | 1.10 | 0.09 | 0.54 |
| Hyperlipidemia to angina | 362 | 1.17 | $2.9 \times 10^{-3}$ | 0.54 |
| Hyperlipidemia to angioplasty | 237 | 1.06 | 0.39 | 0.52 |
| Chronic ischaemic heart disease to CABG | 217 | 1.14 | 0.05 | 0.54 |
| Hypertension to CABG | 121 | 1.19 | 0.06 | 0.55 |
| Angina to CABG | 158 | 1.12 | 0.17 | 0.53 |

**Supplementary Figure 1.**
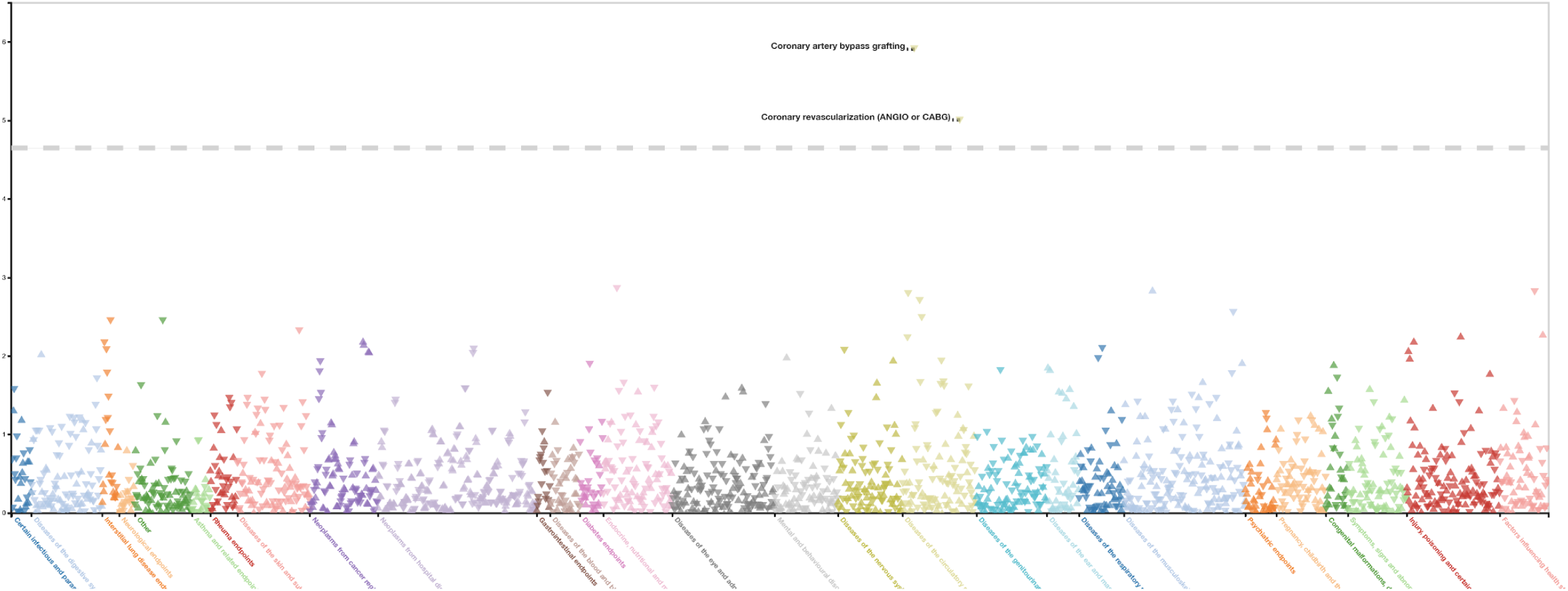
Phewas plot of rs4480535 upstream of *LMO1* shows associates with lower risk of CABG in Finngen. Finngen freeze 4. Variant rs4480535 (11:8302272:A:G) upstream of *LMO1* has a protective effect on coronary artery bypass grafting beta = −0.12, *p* = 1.22×10^-6^. Cases = 4449 and controls = 166928 http://r4.finngen.fi/region/I9_CABG/gene/LMO1

**Supplementary figure 2.**
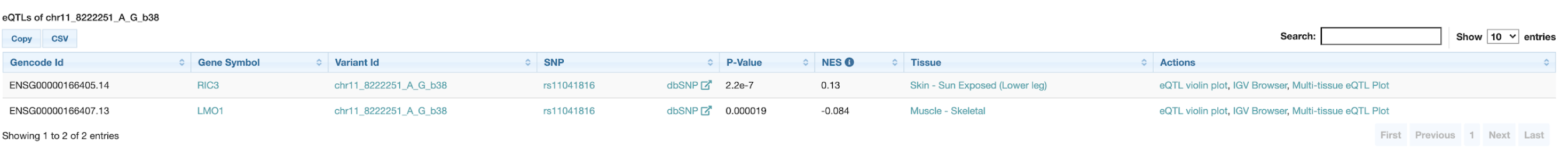
GTEx eQTL for variant rs11041816 identified two genes *RIC3* and *LMO1*

